# Clinical performance of cell free DNA for fetal RhD detection in RhD-negative pregnant individuals in the US

**DOI:** 10.1101/2024.07.24.24310793

**Authors:** Julio F. Mateus Nino, Julia Wynn, Jenny Wiggins-Smith, J. Brett Bryant, J. Kris Citty, J. Kyle Citty, Samir Ahuja, Roger Newman

**Affiliations:** Atrium Health, Charlotte, NC, US; BillionToOne Inc., Menlo Park, CA, US; Medical University of South Carolina, Charleston, SC, US; Shannon Health, San Angelo, TX, US; Unity Health Searcy, Searcy, AR, USA; Wellstar Health System, Marietta, GA, US; University Hospital, Mentor, OH, US

## Abstract

**Objective:** We aimed to evaluate the performance of a cell free DNA (cfDNA) assay that uses next generation sequencing (NGS) with quantitative counting templates (QCT) for the clinical detection of the fetal RHD genotype in a diverse RhD-negative pregnant population in the United States (US).

**Study Design:** This retrospective cohort study was conducted in four US healthcare centers. The same NGS QCT cfDNA fetal RhD assay was offered to non-alloimmunized, RhD-negative pregnant individuals as part of clinical care. Rh immune globulin (RhIG) was administered at the discretion of the provider. The assay’s sensitivity, specificity, and accuracy were calculated considering the neonatal RhD serology results.

**Results:** A total of 401 non-alloimunized RhD-negative pregnancies who received clinical care in the period from August 2020 to November 2023 were included in the analysis. The D antigen cfDNA result was 100% concordant with the neonatal serology, resulting in 100% sensitivity, 100% positive predictive value (both 95% CI: 98.6%-100%), 100% specificity, and 100% negative predictive value (both 95% CI: 97.4%-100%). There were 10 pregnancies where the cfDNA analysis identified a non-RHD gene deletion, including RhDΨ (n=5) and RHD-CE-D hybrid variants (n=5). RhIG was administered to 93% of pregnant individuals with cfDNA results indicating an RhD-positive fetus compared to 75% of pregnant individuals with cfDNA results indicating an RhD-negative fetus, signifying providers were using the results to guide pregnancy management.

**Conclusion:** This cfDNA analysis via NGS for detecting fetal RhD status is highly accurate with no false-positive or false-negative results in 401 racially and ethnically diverse pregnancies with 100% follow up of all live births. This study and prior studies of this assay support a recommendation to offer cfDNA screening for fetal Rh status as an alternative option to prophylactic RhIG for all non-alloimmunized RhD-negative individuals, which will result in more efficient and targeted prenatal care with administration of RhIG only when medically indicated.

## INTRODUCTION

Approximately 15% of pregnant individuals in the United States (US) are RhD-negative.^1,2^ Current national guidelines support the administration of prophylactic anti-D immunoglobulin (RhIG) at 28 weeks of pregnancy and in other circumstances where alloimmunization can occur.^1^ However, in 35-40% of these pregnancies, the fetus is negative for the D antigen and therefore the pregnant person is not at risk for sensitization and RhIG is unnecessary.^2^

Prenatal cell-free DNA (cfDNA) analysis, also known as non-invasive prenatal testing (NIPT), to predict fetal RhD status can be a more efficient and targeted approach to prophylactic RhIG administration in all RhD-negative non-alloimmunzed pregnant patients, limiting the administration of RhIG only to patients carrying an identified RhD-positive fetus. In April 2024, the American College of Obstetrics and Gynecology (ACOG) stated that fetal RhD cfDNA analysis is a reasonable approach for practices experiencing RhIG shortages.^3,4^

The United Kingdom (UK) and several other European countries have used cfDNA analysis to guide the administration of RhIG for over a decade.^5–11^ This approach has not been adopted in the US because of concerns about the inclusivity and accuracy of the European-based assays for the US population.^1^ European assays use polymerase chain reaction (PCR) technology which is particularly impacted by low concentrations of the target gene causing high background noise, DNA amplification variability, and potential for non-specific amplification, leading to inaccurate results particularly at early gestational ages.^12, 13^ Some of these assays have primers that are able to predict the presence a non-RHD gene deletion, genotypes more common in individuals of non-European ancestry; however, they are unable to determine fetal RhD status resulting in a higher frequency of inconclusive results in people of non-European ancestry.^6,12,14–16^ A new cfDNA assay that uses next-generation sequencing (NGS) with quantitative counting template (QCT) technology is currently available in the US.^17^ QCT technology with NGS allows for the precise quantification of DNA at low concentrations. This assay sequences and quantifies of the critical exons of the RHD gene, including those that distinguish it from the RHCE homolog gene and RhDΨ variant, to predict the fetal RhD status for both the RHD-gene deletion and non-RHD-gene deletions at a gestational age as early as 10 weeks. ^17^ Prior studies of this assay in alloimmunized patients demonstrated 100% concordance of the cfDNA results with the neonatal antigen genotype for red blood cell (RBC) antigens including D; however, the clinical use of this assay has not been evaluated in a consecutive cohort of non-alloimmunized pregnant individuals.^17,18^

Given the limitations of European-based assays, particularly for the diverse US population, the objective of this study was to examine the clinical accuracy of NGS QCT cfDNA analysis for the detection of the fetal RHD-genotype for the prediction of the fetal RhD phenotype in a non-alloimmunized, RhD-negative US pregnant population. This study also examined how cfDNA results were used to guide RhIG administration and how the frequency of this changed over the course of the study.

## MATERIALS AND METHODS

This retrospective study was conducted at four healthcare institutions in the US from August 2020 to November 2023, on non-alloimmunized, RhD-negative pregnancies ≥ 10 weeks of gestation, not conceived with an egg donor, or carried by a gestational surrogate with an expected delivery before May 2024 who were having the same cfDNA fetal RhD test (NAME REDACTED) as part of clinical care. All participating institutions and the sponsor site received IRB approval. The study was exempted from patient consent as it was a retrospective study of medical records from clinical care.

As part of standard clinical procedures for the cfDNA fetal RhD assay, the patient blood sample was collected at the provider office and shipped to the central testing laboratory; NAME REDACTED, a CLIA and CAP accredited clinical lab. The methodology and algorithm of the cfDNA fetal RhD laboratory developed test (LDT) did not change during the study and has been described previously in detail.^17^ Briefly, plasma is isolated from a blood sample from the pregnant individual and cfDNA is extracted. Five amplicons across the RHD gene that are unique between the wild-type RHD gene, the RHCE homolog gene, and the RHDΨ variant are amplified and sequenced via NGS. Common polymorphic loci used as reference loci to determine fetal fraction and expected molecular counts for the RHD gene are also amplified and sequenced via NGS. For each amplicon, molecular counts are calculated using QCTs that are added to the sample prior to amplification. The post-amplification reads-per-molecule value is used to compute the absolute detected number of pre-amplification molecules (ADM) and informative polymorphic loci are used to calculate the absolute expected number of molecules (AEM). The number of detected molecules for an antigen of interest, ADM, divided by the number of expected molecules, AEM, is the calibrated fetal RhD antigen fraction (CFAF) for each amplicon. The second highest CFAF across the five amplicons is used as the overall CFAF. A CFAF of 0% is predicted for a D antigen negative fetus and a CFAF of 100% is predicted for a D antigen positive fetus. Only amplification of exon 10 is predicted for an RHD-CE-D hybrid variant associated with an RhD-negative phenotype and amplification of the 37-base pair insertion in exon 4 is expected for the RhDΨ variant. The CFAF value of each amplicon indicates if the fetus, and only the fetus, is negative due to the non-RHD deletion genotype, or the fetus and pregnant person are RhD-negative due to a non-RHD deletion genotype.^17^

The newborn D antigen serology was completed as part of clinical care via the accepted methodology of the clinical institution and results were collected as part of the chart review.

The standard clinical method of type and screen includes forward and reverse typing which includes mixing the RBCs from the patient with RhD antibodies and measuring hemagglutination (forward) and mixing the serum from the patients with RhD antigens and measuring hemagglutination (reverse).^19^ Other abstracted data included pregnant patient demographic information, maternal age, race and ethnicity, and gestational age at the time of testing, results of maternal pregnant person antigen serology, RBC antibody screening, and the frequency and gestational age of antenatal and postnatal RhIG administration. Data were abstracted by research personnel who were not directly informed of the cfDNA results; however, cfDNA results were in the medical records as testing was done as part of clinical care and may have been inadvertently viewed; and therefore, extractors may not have been completely blinded to the prenatal results.

The sensitivity, specificity, positive predictive value (PPV), negative predictive value (NPV), and accuracy of the fetal cfDNA RhD assay were calculated by comparing the predicted fetal RhD status with neonatal RhD serology. Results were considered concordant if cfDNA reported RhD detected, and the neonatal serology was RhD-positive or the cfDNA reported RhD not detected and neonatal serology was RhD-negative. Twin cases were classified as concordant if cfDNA results were RhD detected and neonatal serology for one or both twins was RhD positive, or if cfDNA results were RhD not detected and both twins’ neonatal serology were RhD negative. A competing risk model was used to test for a relationship between fetal loss, and therefore no neonatal serology results, and the cfDNA assay.^20^ A sample size of 335 cfDNA assays was selected on the basis of a conservative predicted sensitivity of 98%, reflective of reported sensitivities of European-based assays and the previously available US assay, to allow the calculation of the assay analytics with a marginal error of 1.5%.^6,12,16^ A two-sample t-test was used to compare the frequency of antenatal RhIG when the cfDNA results reported RhD detected and RhD not detected for all cases. A two-sample t-test was also used to look at the frequency of antenatal RhIG for the cfDNA RhD not detected cases during two time periods: August 2020 and December 2022, when half of the tests occurred, and January 2023 to November 2023, when the second half of the tests occurred. A supplemental meta-analysis of the assay performance was completed by combining the data from the current study with previously published studies of the same assay.^16,17^ Finally, all reported false negatives of this assay received to the laboratory are described in the supplemental results. Analysis was completed in Rv4.2 and 95% confidence intervals were calculated for all metrics.

This investigation met the STROBE (Strengthening the Reporting of Observational Studies in Epidemiology) guidelines for a cohort study.

## Results

There were 410 non-alloimmunized RhD-negative pregnancies in which the cfDNA fetal RhD assay was performed across the four clinical sites during the period studied. The race and ethnicity of the individuals was reflective of the US population (Table 1, Table S1). Pregnancies were excluded if the medical records showed the pregnant person to be RhD-positive or have D-antibodies (were alloimmunized), or the pregnancy was not delivered at the study site. An informative cfDNA fetal RhD result was reported for all 410 pregnancies (no-call rate 0%, 95% CI 0%-0.9%,) without needing a repeat sample. Neonatal serology results were available for the 401 pregnancies that resulted in a live birth. A total of nine pregnancies had a fetal loss related to multiple congenital anomalies (n=2), trisomy 21 (n=1), and unknown reasons (n=6, Figure 1). The competing risk model found no relationship between the cfDNA fetal RhD results and pregnancy outcome (p-value= 0.58, 95% CI 0.98-1.04). Furthermore, the medical record review did not identify a relationship between the pregnancy outcomes and the pregnant person’s RhD status. These cases were excluded from the analysis of the assay performance.

**Figure 1.**
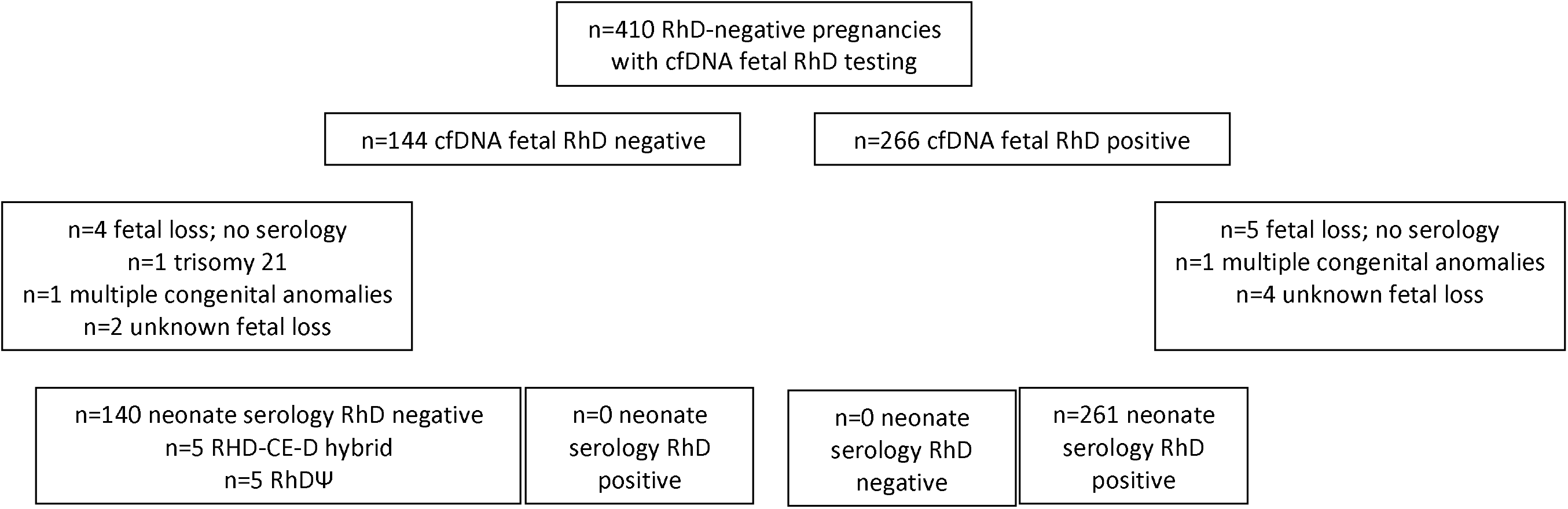
Flow diagram of pregnancies excluded and included in the study.

| **Race and Ethnicity** | **N** | **% of Total** |
| --- | --- | --- |
| Number of Gestations |  |  |
| Singleton | 400 | 99.8% |
| Twin | 1 | 0.2% |
| Maternal Race and Ethnicity |  |  |
| Asian | 3 | 0.8% |
| Black | 31 | 7.8% |
| Hispanic | 68 | 17.1% |
| More than one race | 3 | 0.8% |
| White | 293 | 73.6% |
| Unknown | 3 |  |
|  | **Mean** | **Range** |
| Maternal Age | 28 | 17-45 |
| Fetal Fraction | 8.2% | 1.6%-28.9% |
| Gestational Age At Testing (wk) | 13.82 | 10-34 |
|  | **n** | **%** |
| 1st Trimester | 317 | 79% |
| 2nd Trimester | 80 | 20% |
| 3rd Trimester | 4 | 1% |

Of the 401 cases with neonatal serology, 140 (34.9%) were RhD-negative and 261 (65.1%) were RhD-positive (Figure 1). The predicted fetal D antigen cfDNA result was 100% concordant with the neonatal serology results; resulting in 100% sensitivity (95% CI: 98.6%- 100%), 100% specificity (95% CI: 97.4%-100%), 100% PPV (95% CI: 98.6%-100%), 100% NPV (95% CI: 97.4%-100%), and 100% accuracy (95% CI: 99.1%-100%; Table 2). When the data from the current study and two prior studies were combined, the assay still had 100% performance (Supplemental Results; Table S2). The cfDNA assay also correctly identified the fetal D antigen phenotype in ten cases of non-RHD gene deletions. The predicted fetal D phenotype was concordant with the postnatal D serology phenotype for all of these cases. In five cases, the RhDΨ variant was identified in three patients who identified as Black and two patients who identified as Hispanic (Table S3). The RHD-CE-D hybrid was identified in five other pregnancies including three patients who identified as White, Non-Hispanic, one patient who identified as Black, and one patient who identified as more than one race. In three cases, cfDNA detected the RHD-CE-D hybrid variant in the fetus, but not in the pregnant individual, whereas in two cases the variant was present in the pregnant individual (Table S3).

**Table 2.** Concordance of the cfDNA fetal RhD assay and neonatal D antigen serology and assay performance metrics.

| cfDNA | Neonatal Serology |  |
| --- | --- | --- |
|  | RhD- | RhD+ |
| **RhD Not Detected** | 140 | 0 |
| **RhD Detected** | 0 | 261 |
|  | % | 95% CI |
| **Sensitivity** | 100% | 98.6%-100% |
| **Specificity** | 100% | 97.4%-100% |
| **Positive Predictive Value** | 100% | 98.6%-100% |
| **Negative Predictive Value** | 100% | 97.4%-100% |
| **Accuracy** | 100% | 99.1%-100% |

Of the 616 total doses of RhIG administered in the sample, 364 (59%) doses were administered antenatally including 16 individuals (n=6 with cfDNA RhD not detected results, n=10 with cfDNA RhD detected results) who received two antenatal RhIG doses (Table 3). The frequency of antenatal RhIG administration was significantly higher in pregnancies with the fetal RhD detected results than in those with fetal RhD not detected results (93% vs. 75%; P <0.001). Of the 140 cases with fetal RhD not detected results, 33 did not receive RhIG at any point during care and the other 107 cases received 111 unnecessary RhIG doses (Table 3).

**Table 3.**
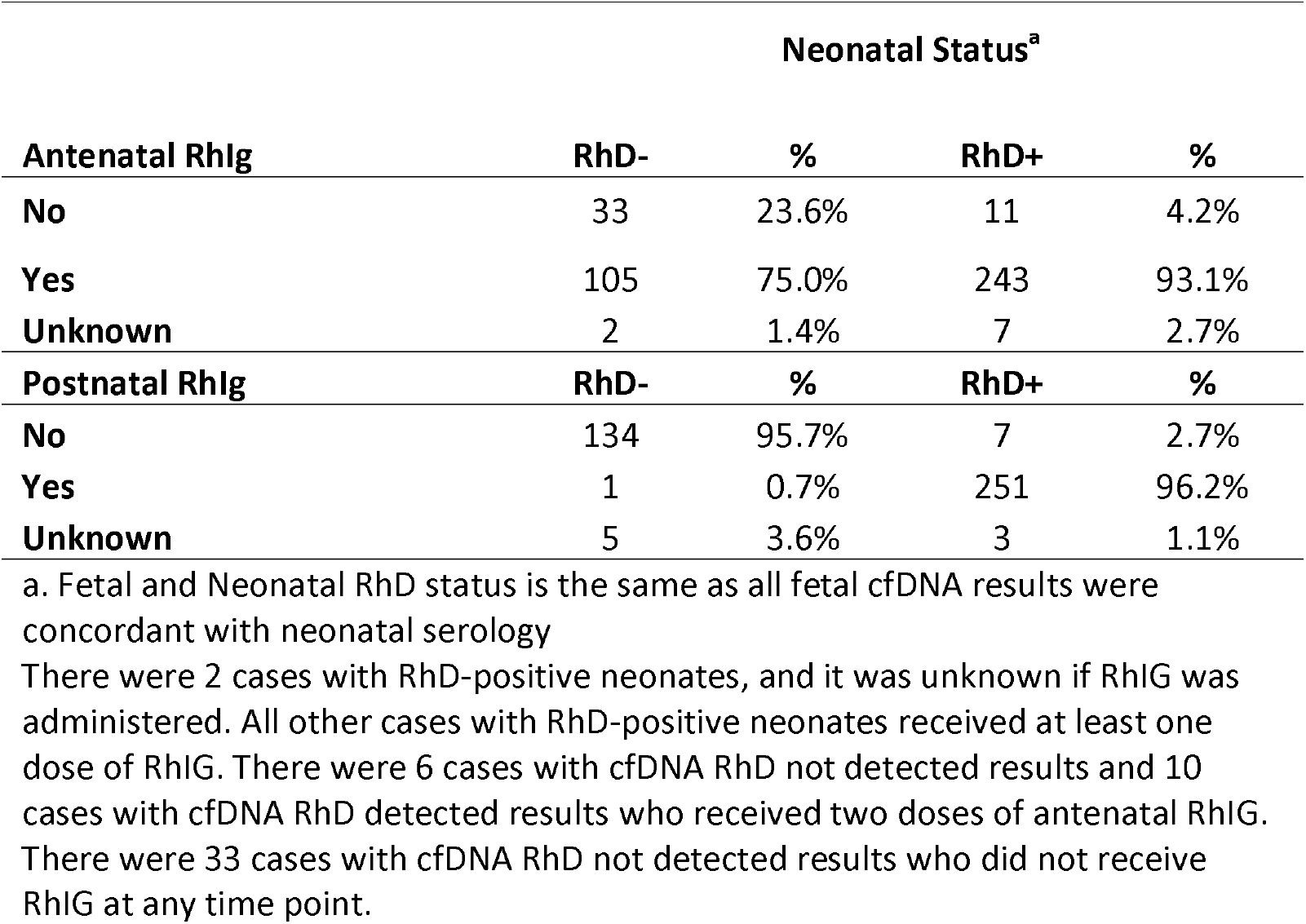
Antenatal and postnatal RhIg administration. A total of 616 doses of RhIG were administered antenatally or postnatally across the 401 cases.

| **Antenatal Rhlg** | **Neonatal Status<sup>a</sup>** |  |  |  |
| --- | --- | --- | --- | --- |
|  | **RhD-** | **%** | **RhD+** | **%** |
| **No** | 33 | 23.6% | 11 | 4.2% |
| **Yes** | 105 | 75.0% | 243 | 93.1% |
| **Unknown** | 2 | 1.4% | 7 | 2.7% |
| **Postnatal Rhlg** | **RhD-** | **%** | **RhD+** | **%** |
| **No** | 134 | 95.7% | 7 | 2.7% |
| **Yes** | 1 | 0.7% | 251 | 96.2% |
| **Unknown** | 5 | 3.6% | 3 | 1.1% |

a. Fetal and Neonatal RhD status is the same as all fetal cfDNA results were concordant with neonatal serology
There were 2 cases with RhD-positive neonates, and it was unknown if Rhlg was administered. All other cases with RhD-positive neonates received at least one dose of Rhlg. There were 6 cases with cfDNA RhD not detected results and 10 cases with cfDNA RhD detected results who received two doses of antenatal Rhlg. There were 33 cases with cfDNA RhD not detected results who did not receive Rhlg at any time point.

When examined by the study site, the difference in RhIG administration based on cfDNA results was significant at three sites (one site had no difference and one site contributed only one case). At one site, no RhIG was administered (antenatally or postnatally) to cases with cfDNA RhD not detected results. Additionally, there was a significant reduction, over time, in RhIG administration in pregnancies with cfDNA fetal RHD not detected results, with 86% of the first 101 pregnancies receiving RhIG versus 69% for next 100 pregnancies s (p-value =0.015)

## DISCUSSION

### Principal Findings

We demonstrated that cfDNA analysis via NGS with QCT for the detection of fetal RhD status is highly accurate in a diverse US clinical population of non-alloimmunized, RhD-negative pregnant individuals. The test performance demonstrated sensitivity, specificity, PPV, NPV, and accuracy of 100%. Importantly, the cfDNA RhD assay was informative in all cases, with a 0% no-call rate, as early as 10 weeks of gestation. Furthermore, the assay correctly identified fetal RhD phenotype in the presence of non RHD-gene deletions. We also observed clinicians using fetal cfDNA RhD results to guide administration of RhIG. Antenatal RhIG was not administered in 33 pregnancies based on fetal cfDNA RhD results without any adverse clinical outcomes.

The results of this study were consistent with prior studies of the performance of this assay. In the first study of the assay, the cfDNA results were concordant with known genotype for all 455 assays using pre-clinical samples (parent/child genomic DNA de-nucleated, sheered to the size of cfDNA and mixed at a concentration proportional to represent a pregnant person and fetal cfDNA ratios) and the neonatal serology for 21 biobanked pregnant person plasma samples.^17^ In a study of 186 alloimmunized pregnancies, including 41 of which were alloimmunized to D, the fetal cfDNA detected antigen genotype was 100% concordant with the neonatal antigen genotype.^18^

Cell-free DNA has been used for over a decade in the UK and European countries to guide the administration of antenatal RhIG.^5–11^ However, European assays use PCR technology, which relies on the assumption that a reference sequence of another gene amplifies at the same rate as the gene of interest resulting in imprecise measurements and requiring larger starting quantities of cfDNA, thereby requiring a later gestational age at the time of testing and resulting in a higher no-call or inconclusive rates, particularly in individuals of non-European ancestry with non-RHD gene deletions.^12^ While some of these assays are able to identify non-RHD gene deletions, due to challenges of PCR technology to quantify DNA molecules of the target sequence at low concentrations, they are not able to determine the fetal RhD status in the presence of the non-RHD gene deletion particularly when the pregnant person has the variant therefore issuing an inconclusive result with a recommendation to administer RhIG. ^5–12^

Additionally, the use of European-based assays for the US population may be logistically complicated and costly. In contrast, the assay examined in this study is NGS-based with QCT technology which allows for the detection and quantification of critical allogeneic exons of the RHD gene.^16,17^ This includes the detection of 37-base pair insertion associated with the RhDΨ variant and differentiation between the RHD gene and the homolog RHCE gene with detection of RHD-CE-D hybrid variants. Quantification with the QCT technology enables accurate fetal RhD prediction at low fetal fractions (early gestational age) with detection of fetal RhD phenotype in the setting of non-RHD gene deletions in both the pregnant person and fetus. In this study the assay predicated fetal RhD status in ten pregnancies with non-RHD gene deletions. In other assays this would have resulted in a 2.5% overall no-call-rate and an 8% no-call-rate among Black and Hispanic individuals in the study sample. Additionally, in this study and prior publications the assay call rate and performance were unaffected by fetal fraction.^16^ Overall, this assay based on NGS and QCT technology is an optimal test for the demographically diverse US pregnant population.

This study showed a lower frequency of antenatal RhIG administration in patients with RhD not detected cfDNA results compared to patients with RhD detected cfDNA results and greater reduction in the frequency of RhIG administration over the course of the study. Overall, the average number of RhIG doses of 1.54 per pregnancy was lower than a prior report in a US population which found 1.80 doses per RhD-negative pregnancy.^21^ Together, these results indicate providers used the assay to guide administration of RhIG even before the change in ACOG recommendations in response to the US shortage and increased the use of the results to guide care overtime.^3,4^

A challenge to cfDNA fetal RhD testing to guide pregnancy management in the US was the availability of a sensitive and cost-effective assay for the population. Previous evaluations of other cfDNA fetal RhD assays for the US population have been shown to be both clinically and economically inferior to a prophylactic RhIG protocol. ^21–23^ This is related to a high rate of inconclusive results leading to unnecessary RhIG administration, lower sensitivity resulting in the potential for an increased frequency of sensitization, and high cost. The current assay has a greater than 99% call rate and sensitivity. It is run on a cost-effective NGS-based platform indicating its potential to have much higher utility and be more cost-effective than prior assays studied. However, a formal US-based health economics study that considers the potential for an ongoing or recurring RhIG shortage may be beneficial. Notably, the implementation of this assay in the UK and other European countries was not based on predicted economic benefits.

Rather, the national adoption in these countries was a multifaceted process based on factors including the clinical performance of the assay for their populations, the reduction in unnecessary medical interventions, and the potential to streamline and improve medical care and neonatal outcomes. ^24^, ^25^

The strengths of this study include the composition of a sample of consecutive pregnancies from the diverse US-based clinics, including over 25% of individuals who identified as non-White, and neonatal serology available for all livebirths. There were ten cases where a non-RHD gene deletion genotype with a predicted RhD-negative phenotype was detected using cfDNA showing the potential of this assay to detect fetal RhD phenotype in the setting of non-RHD gene deletions which are more common in individuals of non-Eurpean ancestry.^14,15^ We also acknowledge the limitations of our study. While the assay correctly determined the fetal RhD status for non-RHD gene deletions, it is a modest sample size. Additionally, as with many prenatal cfDNA assays, this assay is not indicated for pregnant individuals with a history of a bone marrow transplant or solid organ transplant. In these circumstances, the assay is limited due to the potential presence of cfDNA originating from neither the fetus nor the pregnant individual. The assay is also not currently indicated for pregnant individuals with three or more gestations and the current study does not reflect the performance of the assay for twin pregnancies.

In conclusion, these data demonstrate the excellent sensitivity and specificity of this quantitative cfDNA analysis via NGS with QCT technology for detecting fetal RhD status in a diverse US population. These data and the data previously published support offering this assay to non-alloimmunized RhD-negative pregnant individuals in the context of appropriate counseling including risks, benefits and limitations of the test.^17,18^ This implementation will result in more efficient prenatal care and conservation of RhIG by use only in pregnancies where it is medically necessary.

## Data Availability

A complete set of anonymized data is available on request from the corresponding author.

## Acknowledgements

We would like to thank Shannon Rego for her assistance with data analysis and manuscript preparation. We would like to thank individuals who conducted the chart extractions including Todd Morgan, Lindsey Hendry, Arghal Ahmad, Anita K. LaMonica, Becky J. Covington, Brittany Nugent, and Gretchen Hoelscher.

## Conflict of Interest

Dr. Samir Ahuja is a paid consultant from BillionToOne, Inc. Julia Wynn is an employee of BillionToOne, Inc. and has stock options in the company. Drs. Jenny Wiggins-Smith, J. Bret Bryant, J. Kris Citty, J. Kyle Citty and Samir Ahuja received funding for this research project from BillionToOne, Inc.

Precis: cfDNA analysis via NGS is 100% accurate in detecting fetal RhD status in 401 racially and ethnically diverse pregnancies with 100% follow up of all live births.

## Author contributions

JFMN contributed conceptualization, data curation, writing the original draft and review and edition the manuscript. JW contributed to conceptualization, project administration, formal analysis and review and editing of the manuscript. JWS, JBB, KC, KC, and SA contributed to data curation, supervision and review and editing of the manuscript. RN contributed to formal review and editing of the manuscript.

## Supplemental Results

The race and ethnicity breakdown of the sample is reflective of the US population and the frequency of the Rh-negative blood type in these race and ethnic groups.^1^ Approximately 14- 16% of the US population is RhD-negative but this phenotype is more common in White, non-Hispanic (∼17.3%) and less common in Asian individuals (∼1.7%) and Black individuals (∼7.1%).^2^ Based on these frequencies, we would expect approximately 28 Black individuals and 3 Asian individuals in a sample of 410 RhD negative pregnant individuals from the US population; consistent with the frequencies observed in this study (Table S1).

**Supplemental Table 1.** Published blood type, race and ethnicity frequencies for the US population with the expected and observed number of each race and ethnicity for a sample of 410 US RhD-negative individuals. The chi-square statistic is 5.78 (excluding more than one race and unknown). The p-value is 0.12.

|  | US Frequency |  |  |  |
| --- | --- | --- | --- | --- |
|  | All | RhD-Negative | Expected | Observed |
| Asian | 6.4% | 1.7% | 3 | 3 |
| Black | 13.7% | 7.1% | 30 | 31 |
| Hispanic | 19.5% | 7.2% | 44 | 68 |
| White | 58.4% | 17.3% | 313 | 293 |
| More than one Race | 2.0% | unknown | unknown | 3 |
| Unknown | N/A | N/A | N/A | 3 |

This study and two other published studies have examined the performance of the same cfDNA RhD assay. Together, the performance of 925 RhD cfDNA assays has been studied. This includes 470 clinical samples from RhD-negative pregnant individuals and 465 pre-clinical samples (parent/child DNA sheared to the size of cfDNA and mixed at concentration proportions to represent a pregnant person and fetal cfDNA ratios).^3,4^ Across all 925 assays, the cfDNA results were concordant with the known serology and/or genotype of the resulting neonate. This combined cohort had a sensitivity and PPV of 100% (95% CI 99.3%-100%) and specificity and NPV of 100% (95% CI 98.9%-100%; Table S2)

**Supplemental Table 2.** Concordance of the cfDNA fetal RhD assay and neonatal RhD genotype or serology across three studies.

| cfDNA | Neonatal Serology |  |
| --- | --- | --- |
|  | RhD- | RhD+ |
| **RhD Not Detected** | 572 | 0 |
| **RhD Detected** | 0 | 353 |
|  | **%** | **95% CI** |
| **Sensitivity** | 100% | 99.30% to 100.00% |
| **Specificity** | 100% | 98.89% to 100.00% |
| **Positive Predictive Value** | 100% | 99.30% to 100.00% |
| **Negative Predictive Value** | 100% | 98.89% to 100.00% |
| **Accuracy** | 100% | 99.57% to 100.00% |

Additionally, the clinical laboratory has completed over 10,000 RhD cfDNA assays in RhD-negative pregnant individuals. Across all clinical samples tested at our laboratory, there have been two reported false negatives with complete follow up. In both cases, the provider reported the neonate tested RhD-positive at birth however follow-up testing of the neonate showed RhD-negative. Therefore, there are presently no confirmed false negative cases.

The frequency of non-RHD gene deletion variants in Black (13%), Hispanic (3%) and White, non-Hispanic (1%) RhD-negative individuals was consistent with the reported frequencies of non-RHD gene deletion variants in these racial and ethnic groups (Table S3).^5^

**Supplemental Table 3.**
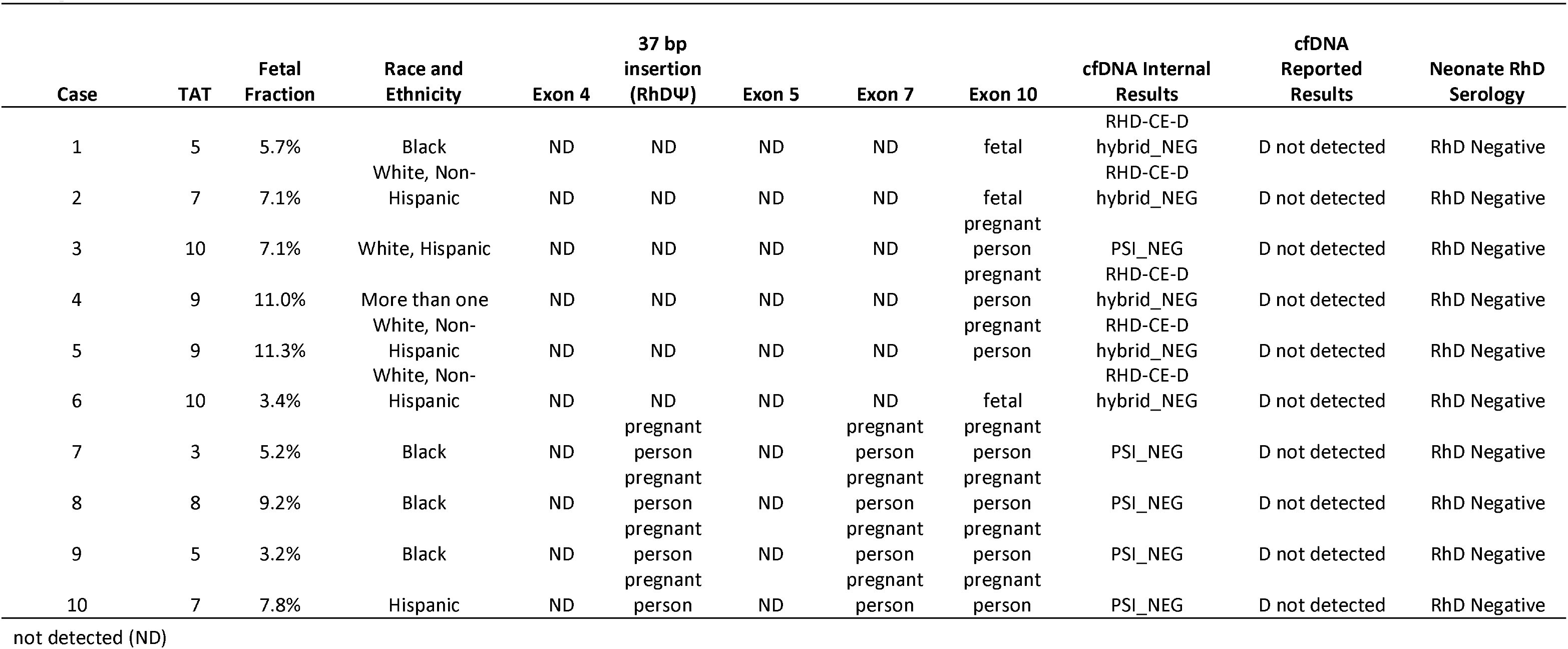
Demographics and cfDNA predicted genotype and phenotype for the 10 cases where a non-RHD gene deletion was detected by cfDNA. The data includes whether or not the amplicon for the exon was detected and if it was at a quantity suggestive for fetal and/or maternal origin. In an RhD-negative person with the RhDΨ variant only this variant would be present without any other RHD exons. In a RhD-negative person with the RHD-CE-D hybrid variant, only RHD exon 10 is present. All other exons are from the RHCE gene.

